# Pre-trained convolutional neural network with transfer learning by artificial illustrated images classify power Doppler ultrasound images of rheumatoid arthritis joints

**DOI:** 10.1101/2024.08.30.24312848

**Authors:** Jun Fukae, Yoshiharu Amasaki, Yuichiro Fujieda, Yuki Sone, Ken Katagishi, Tatsunori Horie, Tamotsu Kamishima, Tatsuya Atsumi

## Abstract

**Objectives:** To study the classification performance of a pre-trained convolutional neural network (CNN) with transfer learning by artificial images of the joint ultrasonography in rheumatoid arthritis (RA).

**Methods:** We focused on abnormal synovial vascularity and created 870 artificial ultrasound joint images based on the European League Against Rheumatism/Outcome Measure in Rheumatology scoring system. One CNN, the Visual Geometry Group (VGG)-16 was trained with transfer learning using the 870 artificial images for initial training and the original plus five additional images for second training. Actual joint ultrasound images obtained from patients with RA were used for testing our models.

**Results:** We obtained 156 actual ultrasound joint images from 74 patients with RA. Our initial model showed moderate classification performance, but grade 1 was especially low (area under curve (AUC) 0.59). In our second model, grade 1 showed improvement (AUC 0.73).

**Conclusions:** We concluded that artificial images were useful for training VGG-16. Our novel approach of using artificial images as an alternative to actual images for training CNN has the potential to be applied in medical imaging fields that face difficulties in collecting real clinical images.

**Registration of clinical trials:** This study was registered in UMIN Clinical Trials Registry (UMIN000054321).

## INTRODUCTION

In the past several years, artificial intelligence (AI) has made remarkable progress in image recognition. AI has also progressed in the medical imaging field such that it can now classify clinical images and detect disease abnormalities (1,2). As AI advances, it requires a number of actual clinical images for training to obtain these abilities, and construction of datasets of medical images is currently underway all over the world. However, the collection of actual clinical images is difficult because it involves issues of patient personal privacy.

In rheumatoid arthritis (RA), joint ultrasonography is essential for evaluating the arthritis. Semi-quantitative 4-grade scoring was defined to measure abnormal synovial vascularity by the European League Against Rheumatism/Outcome Measure in Rheumatology (EULAR/OMERACT) (3). The scoring showed significantly fine inter-rater agreement among multiple human raters, but the raters needed clinical training and experience to become expert (4). Contrastingly, AI does not require much time to learn. We devised an idea to use artificial images instead of actual clinical images for training AI. Artificial images are not subject to personal privacy problems and are easy to create. As one of the AI technologies, deep learning mimics brain neural networks via mathematical models. Convolutional neural networks (CNNs) are a representative image recognition technology and one aspect of deep learning. In this study, we evaluated a CNN that was trained by using artificial joint ultrasound images to classify actual ultrasound joint images of patients with RA.

## METHODS

### Convolutional neural network

We used the Visual Geometry Group (VGG)-16 by MATLAB^®^ software R2024a (MathWorks, Inc., Natick, MA) in this study. VGG-16 has 13 convolutional layers and is often used in research for medical imaging classification (5). VGG-16 was pre-trained by the ImageNet database (https://www.image-net.org/index.php), which contained more than one million variable images and could classify 1000 objects. In this study, the pre-trained VGG-16 was further enhanced with transfer learning using our artificial images. The architecture of our models, we removed the last three layers of VGG-16 and replaced them with a fully connected layer, a dropout layer, a softmax layer, and a classification layer. The fully connected layer had learning rate coefficient set to ten times for both weights and biases and applied L2 regularisation. The dropout layer randomly deactivated 30% of the nodes. The softmax and classification layers were adapted for a new four-class classification. We carefully tuned the training process and minimised overfitting. The parameters of transfer learning are described below. The maximum number of epochs was set to 11, the batch size was 32, and the learning rate was set as 0.0001. Data were shuffled before each epoch, and the validation data were set to supply every 30 epochs. Data augmentation was added to reduce the risk of overfitting.

### Creating training data for the convolutional neural network

Seventeen basic ultrasound joint images were created by using CLIP STUDIO PAINT^®^ illustration software 3.02 (CELSYS, Tokyo, JAPAN) by a rheumatologist (JF). The rheumatologist (JF) drew images by hand using a pen tablet Wacom Intuos S^®^ (Wacom, Saitama, JAPAN). These basic images were drawn as dorsal scanning images of metacarpophalangeal (MCP) joints showing abnormal synovial vascularity according to the EULAR/OMERACT scoring system (grade 0: 5 images, grade 1: 4 images, grade 2: 4 images, and grade 3: 4 images). Synovial vascularity was drawn as an aggregation of red pixel dots that localised in the joint capsule. Structure alteration such as bone erosion was not included in the images. Representative basic images are shown in Supplementary Figure 1. Each basic image contains several imaging layers that include joint tissue, bone surface and vascular images, respectively. At least one modification such as object moving and scaling was added to the imaging layers of each basic image and then all layers were combined to generate one image by a rheumatologist (JF) or modifier (YFuk) (Supplementary Figure 2). All images were confirmed by the rheumatologist (JF) as to whether they followed scoring rules. In total, 870 images were obtained (grade 0: 212 images, grade 1: 236 images, grade 2: 222 images, and grade 3: 200 images).

### Actual clinical images for testing performance of the convolutional neural network

In this study, actual images of joint ultrasonography were retrospectively obtained from 74 patients with RA who underwent examinations at Kuriyama Red Cross Hospital. All the patients satisfied the 2010 American College of Rheumatology (ACR)/EULAR classification criteria for RA (6). The characteristics of the patients are shown in Supplementary Table 1. In this study, 156 images of MCP joint ultrasonography were registered. Two expert raters (KK, TH) evaluated synovial vascularity according to the EULAR/OMERACT scoring system.

The weighted kappa value of inter-rater agreement among the two raters was 0.80 (95% confidence interval (CI) 0.73 to 0.86). A third expert rater (YFuj) double-blindly evaluated the 29 images whose score differed between the first two raters. Among the results of the three raters, we used the images whose score matched more than two raters to test our model (grade 0: 43 images, grade 1: 41 images, grade 2: 67 images, and grade 3: 5 images).

### Creating additional training data for the convolutional neural network

We trained VGG-16 with the data created and then tested it with actual clinical images as described above. Our initial model was unable to classify some clinical images correctly (Table 1). Therefore, we focused on and randomly chose misclassified images that were truly grade 1 but that were classified as grade 2 and then created additional artificial images. Five artificial images were created from each of the misclassified images. Each artificial image was modelled after the original misclassified image. We changed the training data by adding the additional artificial images and then found that classification power for some cases rather worsened (data not shown). Finally, five artificial images based on one misclassified image were selected. The VGG-16 was newly trained with transfer learning as a second model by combining the original plus the five artificial images.

**Table 1.**
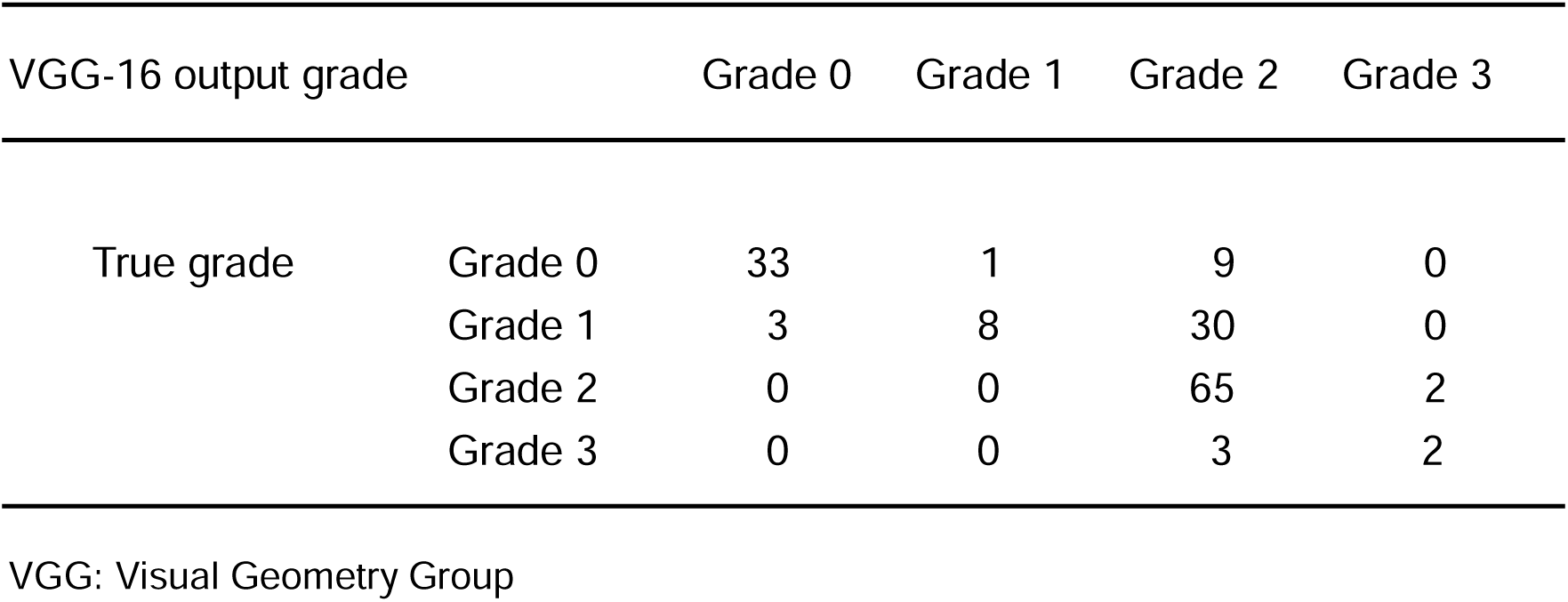
Confusion matrix of the initial model.

### The progress of our study

In our study, artificial images were used for training VGG-16 to establish the initial model, and then the initial model classified actual clinical images. We performed error analysis on the misclassified images, and then created additional images. Original plus additional images were used for training VGG-16 to establish the second model. Flow chart is shown in Figure 1.

**Figure 1.**
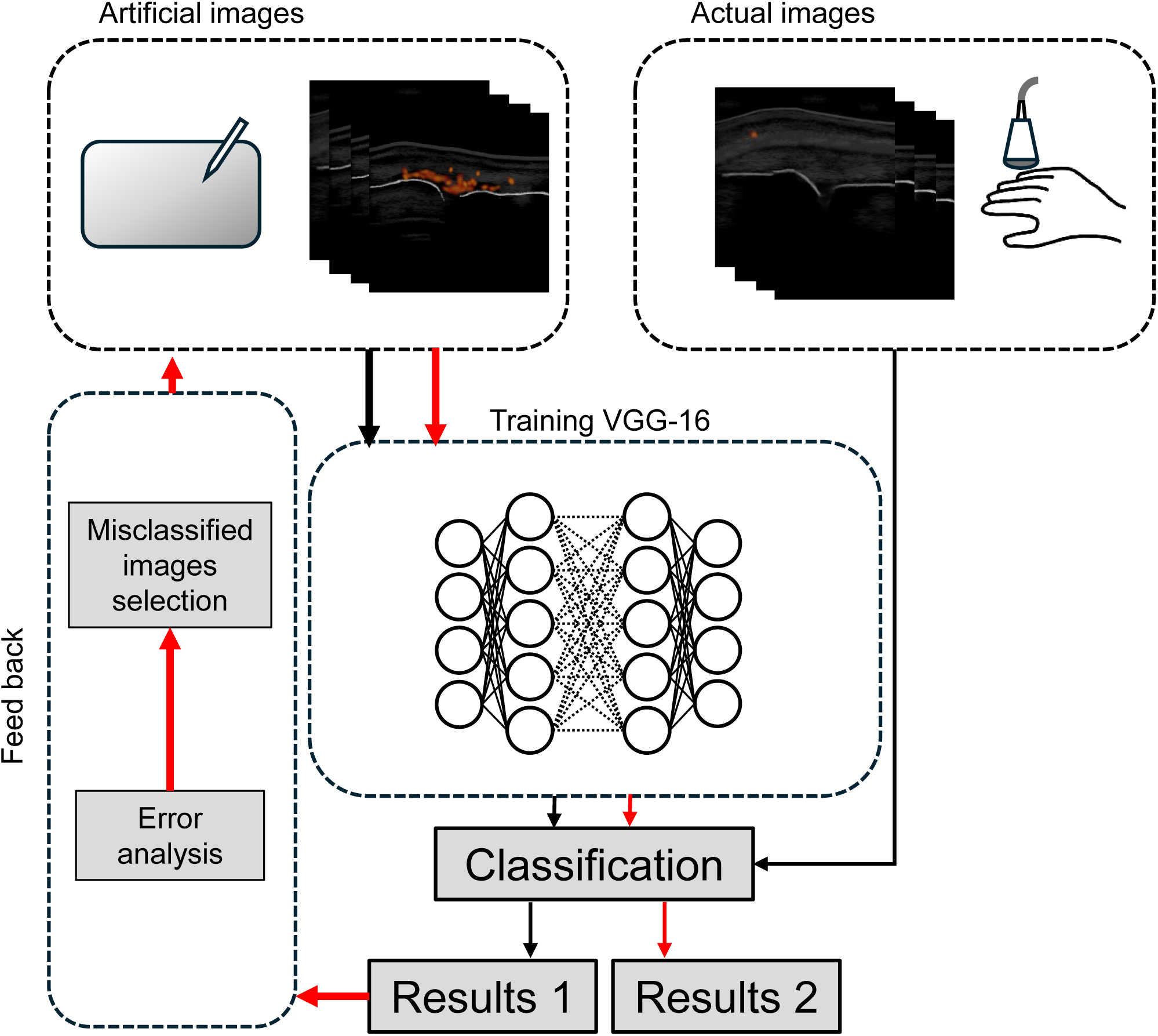
Flow chart of our study. The study proceeded as follows: artificial images were used for training Visual Geometry Group (VGG)-16 to establish the initial model, and then classification of actual images was performed (black line). Original plus additional images were used for training to establish the second model, and then classification of actual images was performed (red line).

### The cross-validation for the second model

To assess the model’s performance and generalizability of the second model, a five-fold cross-validation was performed. The training data was split into five parts and trained five times. Using a different fold as the validation and the remaining as the training. The mean accuracy across the five cross-validation runs was calculated.

### Statistics analysis

Indexes of accuracy, precision, recall, F-score, and specificity were calculated to evaluate CNN performance of each grade classification. The F-score was the harmonic mean of precision and recall ranging from 0 to 1 (approaching 1 meant good balance between precision and recall). Receiver Operating Characteristic (ROC) curve was shown as a trade-off between sensitivity (or recall) and specificity and area under curve (AUC) of each ROC curve was calculated. Confusion matrices for multiclass classification were used to show the whole picture of grade classification. Inter-rater agreement among the two raters was analysed according to the weighted kappa value, which approaches 1 as concordance becomes stronger (0.61-0.80 is considered to be good, and >0.8 is considered to be very good). Statistical analyses were performed with the use of Excel^®^ for Microsoft 365 MSO 2404 (Microsoft, Redmond, WA) and MedCalc^®^ 22.023 (MedCalc Software, Mariakerke, Belgium).

### Ethical approval

This study was approved by the local ethics committees of Kuriyama Red Cross Hospital. The study outline was published on the hospital’s homepage at https://kuriyama.jrc.or.jp/outpatient/, and an opt-out management strategy was used to collect the patient information. All the patients included in the study were above 18 years of age. This study was conducted in accordance with the Declaration of Helsinki. This study reported according to the STARD (Standard for Reporting of Diagnostic Accuracy Studies) guidelines.

## RESULTS

In our VGG-16 with transfer learning by artificial images, the initial model showed the following representative results for three trials of independent learning. The respective values for accuracy, precision, recall, F-score, specificity, and AUC for the four grades were grade 0: 0.92, 0.92, 0.77, 0.84, 0.97, and 0.87; grade 1: 0.78, 0.89, 0.20, 0.32, 0.99, and 0.59; grade 2: 0.72, 0.61, 0.97, 0.75, 0.53, and 0.75; and grade 3: 0.97, 0.5, 0.4, 0.44, 0.99, and 0.69. The confusion matrix for multiclass classification for the initial model is shown in Table 1. The ROC curve of the initial model of each grade is shown in Figure 2. The weighted kappa value of inter-rater agreement between the true grades and the initial model’s output was 0.62 (95% Confidence interval (CI) 0.52 to 0.71).

**Figure 2.**
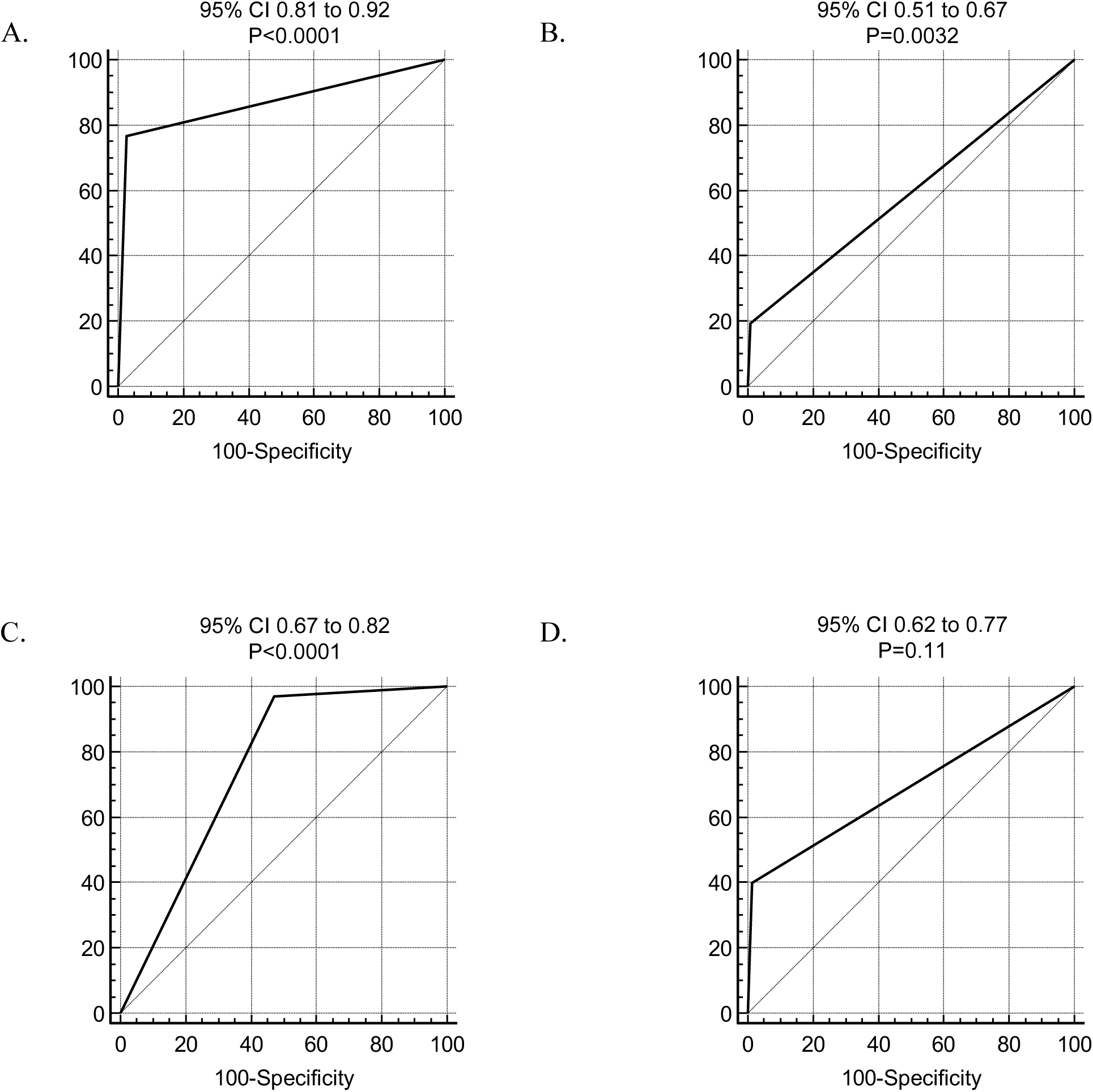
Receiver Operating Characteristic (ROC) curves of the initial model. Four ROC curves were shown. A. Grade 0, B. Grade 1, C. Grade 2, D. Grade 3. CI: Confidence interval

We next trained the VGG-16 CNN with transfer learning by combining the original plus additional training data. This second model showed the following representative results for three trials of independent learning. The respective values for accuracy, precision, recall, F-score, specificity, and AUC were grade 0: 0.90, 0.80, 0.86, 0.83, 0.92, and 0.89; grade 1: 0.83, 0.75, 0.51, 0.61, 0.94, and 0.73; Grade 2: 0.80, 0.73, 0.87, 0.79, 0.75, and 0.81; and grade 3: 0.97, 0.5, 0.2, 0.29, 0.99, and 0.60. The confusion matrix for multiclass classification for the second model is shown in Table 2. The ROC curve of the second model of each grade is shown in Figure 3. The weighted kappa value of inter-rater agreement between the true grades and the second model’s output was 0.69 (95% CI 0.60 to 0.78). The mean accuracy of second model cross-validation was 0.99543.

**Figure 3.**
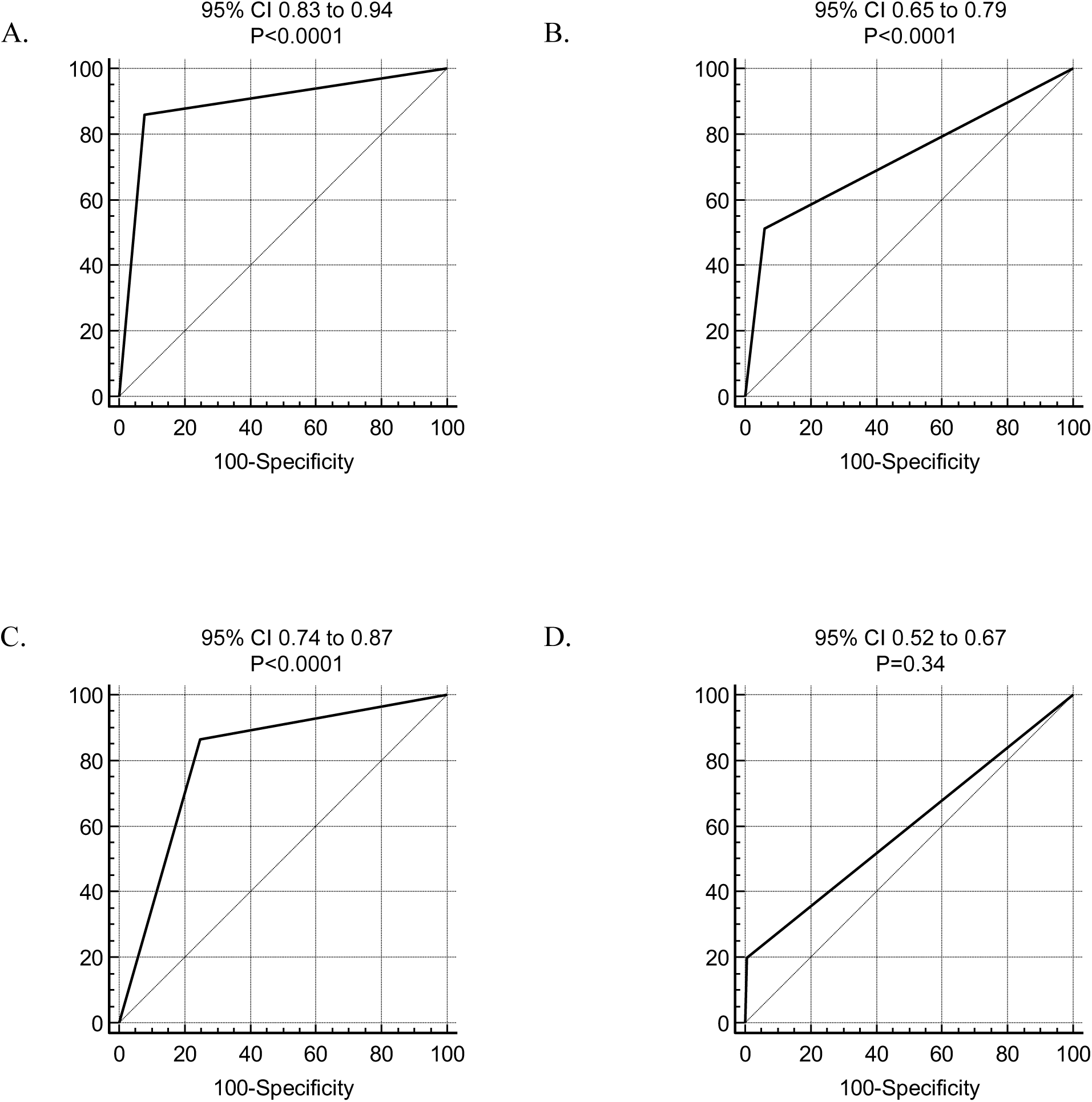
Receiver Operating Characteristic (ROC) curves of the secondary model. Four ROC curves were shown. A. Grade 0, B. Grade 1, C. Grade 2, D. Grade 3. CI: Confidence interval

**Table 2.**
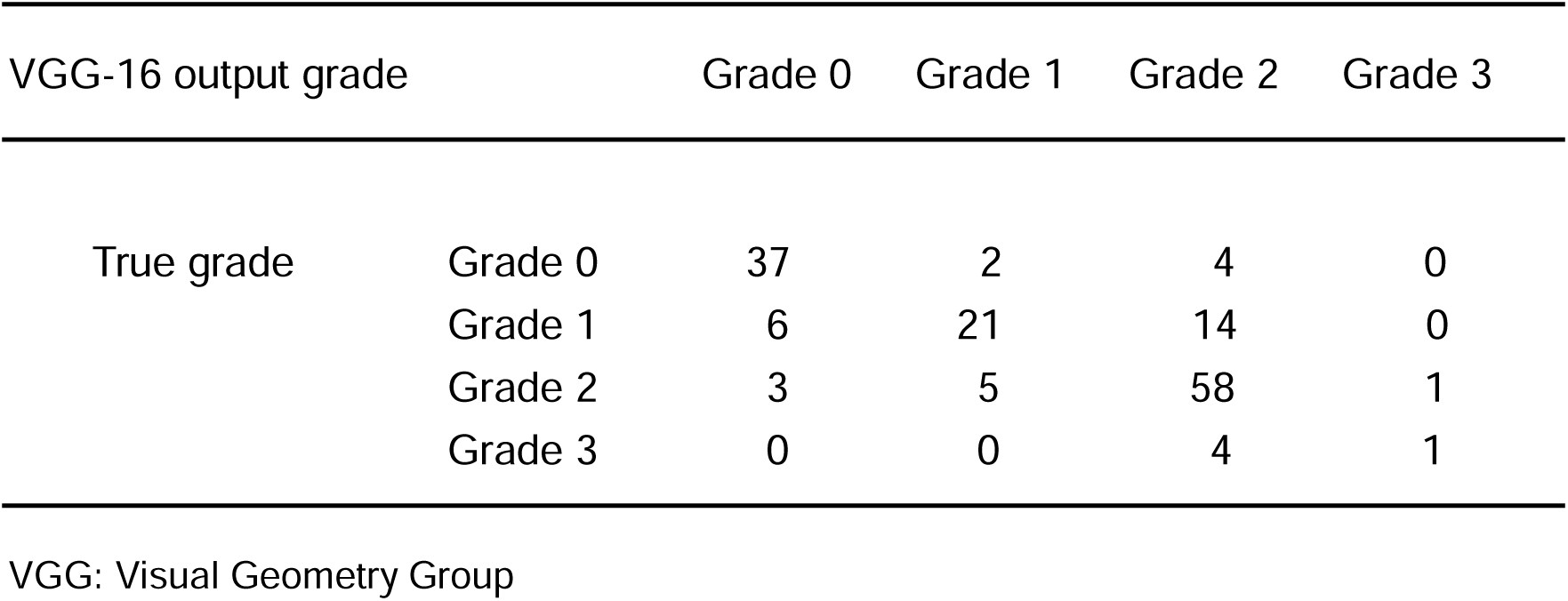
Confusion matrix of the secondary model.

| VGG-16 output grade |  | Grade 0 | Grade 1 | Grade 2 | Grade 3 |
| --- | --- | --- | --- | --- | --- |
| True grade | Grade 0 | 37 | 2 | 4 | 0 |
|  | Grade 1 | 6 | 21 | 14 | 0 |
|  | Grade 2 | 3 | 5 | 58 | 1 |
|  | Grade 3 | 0 | 0 | 4 | 1 |

VGG: Visual Geometry Group

## DISCUSSION

This study was a novel attempt at using artificial images that were created by digital illustration for training of a medical CNN in the field of rheumatology. Our results showed that a pre-trained VGG-16 with transfer learning using artificial illustrated images could classify actual ultrasound joint images. Our models showed moderate classification ability and good inter-rater agreement with human raters. The practical application of our models could lead to more consistent and objective assessment in clinical trials and practice.

Traditionally, real-world clinical images have been used to train CNNs in the medical field. The reasons for using actual images were based on the idea that medical images were precise depictions of normal and abnormal conditions, and medical CNNs could not learn pathological features without using actual images. In previous studies on joint ultrasonography in RA, several groups have studied the classification of joint images by using CNNs (7–10). They used actual clinical images to train the CNNs and reported good results in their studies. One of them, Christensen et al. used 1678 actual ultrasound images for training CNN. Their model achieved an overall four class accuracy of 0.839 in the EULAR/OMERACT synovitis score classification (9). However, it was not easy to collect a large number of clinical images, and there were various problems in collecting training data related to data transparency and privacy protection. These issues were strongly dependent on local and international laws and regulations.

The basic artificial joint images that we used in this study were illustrated by an experienced rheumatologist JF and were added to with modifications by JF and YFuk to increase the number of training images. The process proceeded with ease and required no specialised illustration skills or deep medical knowledge. In contrast to actual images, the artificial images cause no difficulty in collection, and no privacy problems exist. In our study, the confusion matrix (Table 1) of our initial model revealed that classification power especially for grade 1 was inferior to that of the other grades. We analysed the training data and then focused on images of grade 1 that were misclassified as grade 2. We created additional training images as explained in the Methods section. A second model trained by the original plus additional training images improved the classification power (Table 2). For this additional data, we randomly selected several grade 1 images misclassified as grade 2 and then created 5 images from each of them. When adding these images for training, there were some cases that rather worsened the classification results. Therefore, through trial and error, we checked and finally selected useful images. Our method of using artificial images has the advantages of increasing the amount of available training data and responding to misclassified problems.

As for the second model, there was a risk of data leakage, we checked training data splitting, data preprocessing independence and then performed cross-validation. Our second model showed high cross-validation accuracy and moderate classification performance to the actual images. We next removed the original image which was basic for additional 5 images from testing data then performed classification. The whole picture of grade classification showed the same result (data not shown). From this, we concluded the risk of the data leakage was low in our second model. Further validation will be necessary to confirm the generalizability of our model.

Abnormal synovial vascularity was focused on in this study. Power Doppler ultrasound rendered the vascular image in red with a background of black and grey, so VGG-16 might easily focus on it. In the pathology of abnormal synovial vascularity of RA, it is well known that blood flow increases as the inflammation worsens. EULAR/OMERACT defined the degree of synovial vascularity as a simple semi-quantitative 4-grade score. Thus, it was easy to reproduce abnormal synovial vascularity of the four grades in the artificial images.

In the medical image for deep learning study, generative adversarial networks (GANs) were focused on because of their precision image generation ability (11). Although our artificial images were not finely detailed, they were useful as training data. Our novel approach of using artificial images for training CNN as an alternative to actual images has the potential to be applied in medical imaging fields that face difficulties in collecting real clinical images. However, further research will be necessary to confirm this.

There are limitations regarding our results. The number of actual images for grade 3 was small. The sample size of grade 3 images needs to increase to certify reliability. The artificial images that we created were more monotonous and less varied in detail than actual images, and therefore, our model might respond to some cases but not to all. Nonetheless, our methods of creating additional training images were easy and allowed rapid correction of misclassification errors. In our method, feedback of the error analysis linked to training data diversification and improvement of model performance. However, there is always a risk for data leakage or overfitting, therefore the feedback process needs to proceed with caution.

Medical professionals accumulate experience in examining actual images and also learn pathology with schematic images. We hypothesise that CNNs may increase the performance of medical image classification when they learn with both actual and artificial images. This is a topic for future research.

Considering the potential of CNNs, they were originally developed as technology for detecting imaging features by mathematical processing, and they sometimes discovered new features or focus points that were previously unknown, and thus, analysis by this method was important. The feedback from new findings of AI might provide new knowledge to humans. Creating training data based on new knowledge and then using them for training AI might expand its potential. Humans and AI each should have a positive influence on the other. In the medical field, progress in AI should be supportive, but not suppressive, to expand the potential of human clinicians.

## Supporting information

Suppementary Figure 1.

## Data Availability

The training data for initial and secondary models available from the link below on reasonable request.
https://center6.umin.ac.jp/cgi-bin/ctr_e/ctr_view.cgi?recptno=R000061975

## Author contributions

Study conception and design. JF, YA, YFuj, TK, TA Data preparation and acquisition. JF, YA, YFuk, YS, KK Data analysis. JF, YA, KK, TH, YFuj Manuscript preparation. JF, YA, YFuj, TK, TA

## Acknowledgement

We thank Yusuke Fukae for generating training data as an image modifier.

---

**Supplementary Figure 1.** Basic artificial images of joint ultrasonography. Four representative images from 17 basic images of joint ultrasonography are illustrated as dorsal scanning of metacarpophalangeal joints showing abnormal synovial vascularity according to the European League Against Rheumatism/Outcome Measure in Rheumatology scoring system. A. Grade 0, B. Grade 1, C. grade 2, and D. Grade 3.

**Supplementary Figure 2.** Generation of training images. One or more modifications were applied to the image layers, which were then combined to generate new artificial images. The left side of the figure shows the image layers, each of which illustrates joint tissue, bone, and synovial vascularity, respectively. The right side shows the artificial image that was generated after combining the images on the left.

**Supplementary Table 1.**
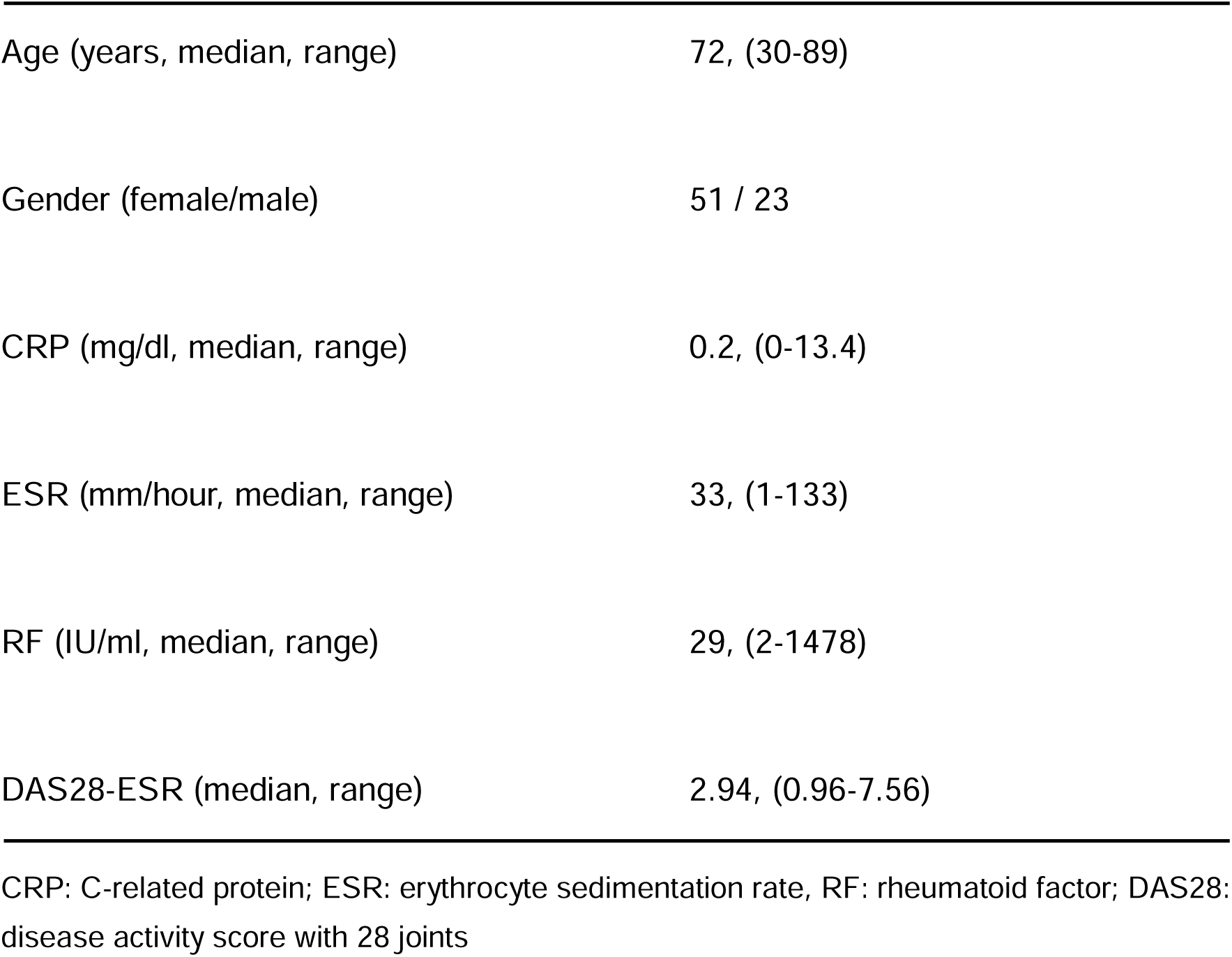
Characteristics of the patients with rheumatoid arthritis.

|  |  |
| --- | --- |
| Age (years, median, range) | 72, (30-89) |
| Gender (female/male) | 51 / 23 |
| CRP (mg/dl, median, range) | 0.2, (0-13.4) |
| ESR (mm/hour, median, range) | 33, (1-133) |
| RF (IU/ml, median, range) | 29, (2-1478) |
| DAS28-ESR (median, range) | 2.94, (0.96-7.56) |

CRP: C-related protein; ESR: erythrocyte sedimentation rate, RF: rheumatoid factor; DAS28: disease activity score with 28 joints

