## Supplementary figures and images for "Pre-trained convolutional neural network with transfer learning by artificial illustrated images classify power Doppler ultrasound images of rheumatoid arthritis joints"

### Suppementary Figure 1.

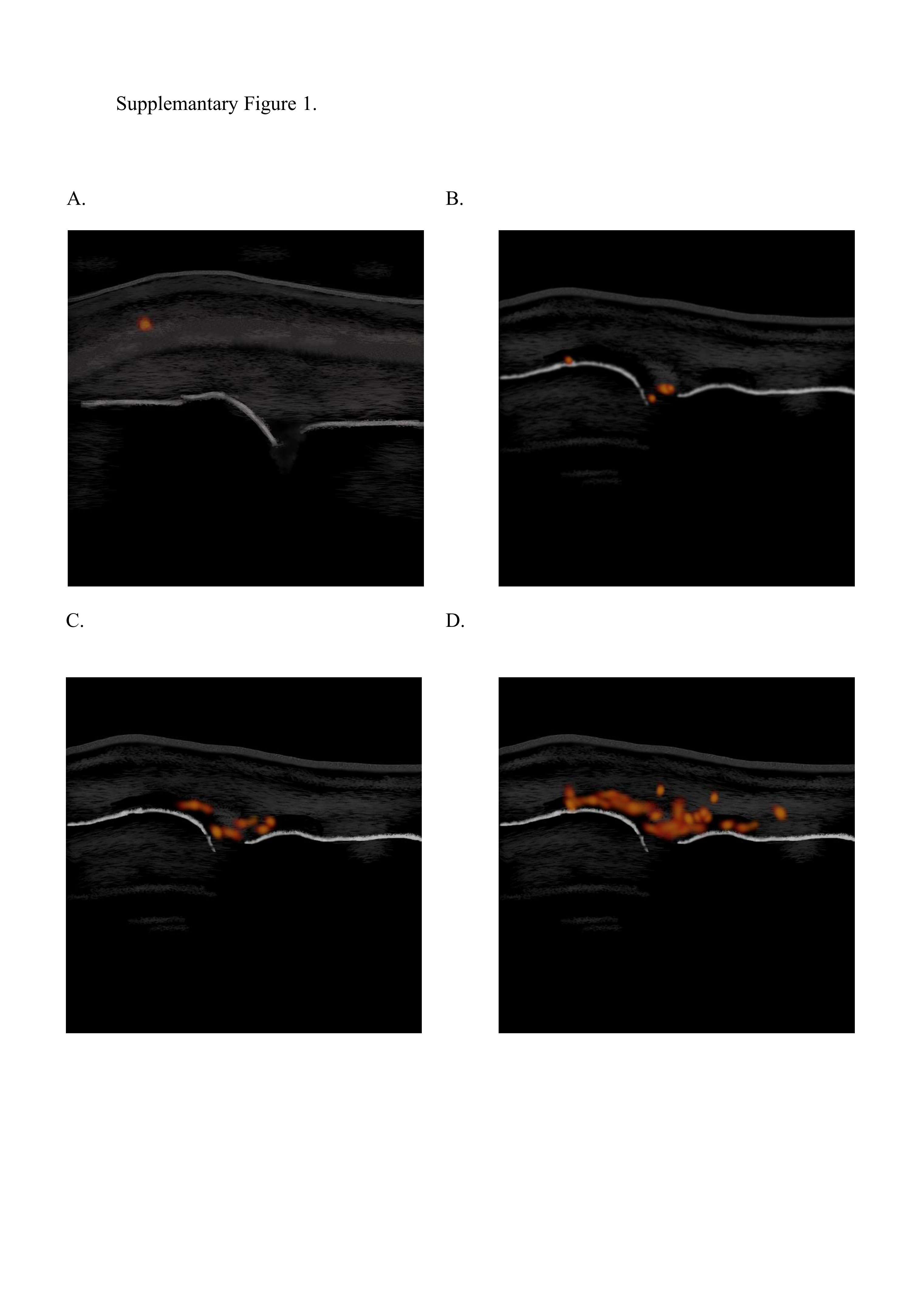

### Supplementary Figure 2.

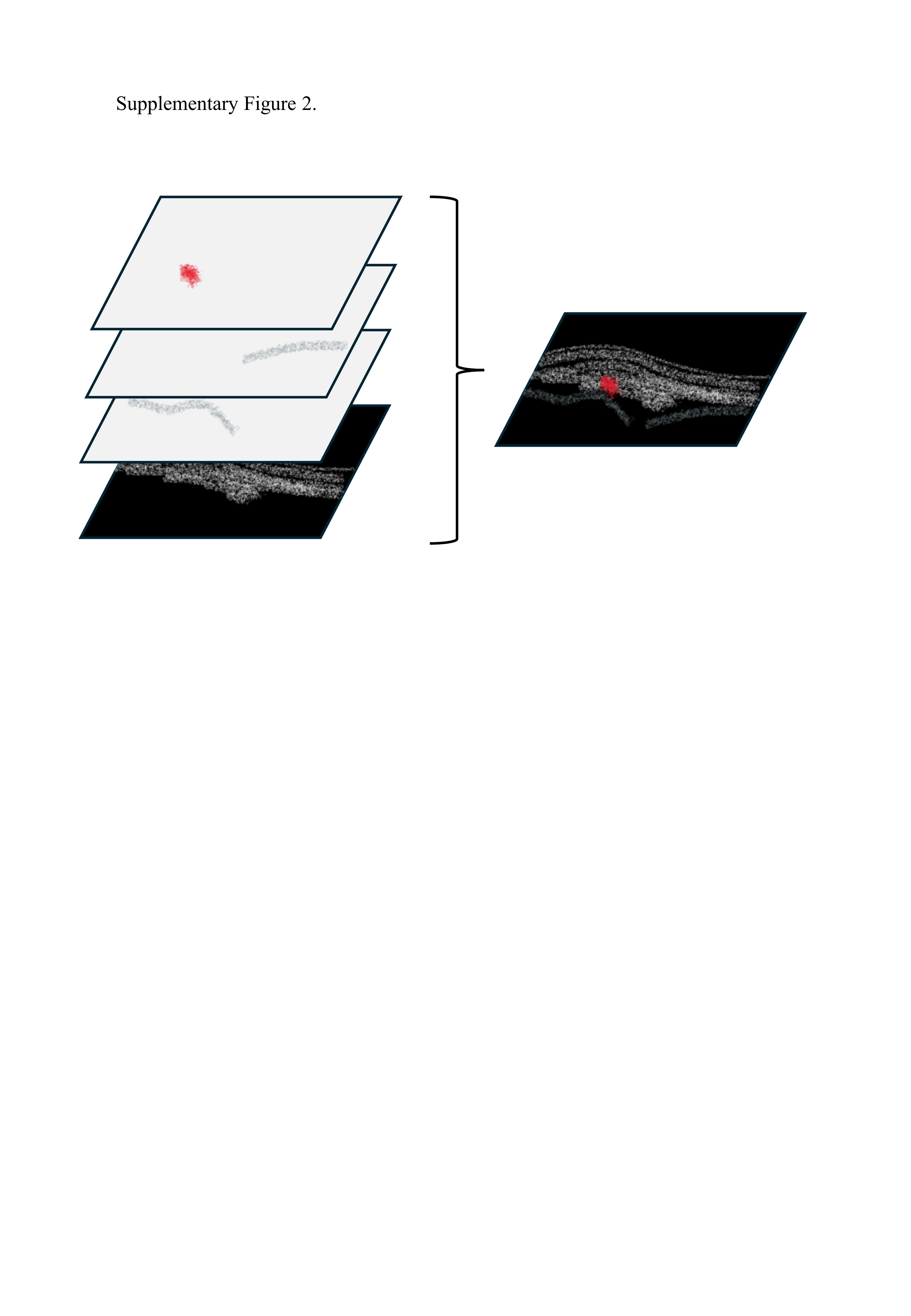
